# Adherence to Chemotherapy Among Patients with Advanced Epithelial Ovarian Cancer in the Netherlands and Its Impact on Survival: A Nationwide Cohort Study

**DOI:** 10.64898/2026.01.30.26345198

**Authors:** S.A. Said, H.H.B. Wenzel, A.M. van Altena, J.E.W. Walraven, J. IntHout, J.A. de Hullu, M.A. van der Aa

## Abstract

**Objective:** Population-based information regarding adherence to first-line chemotherapy in epithelial ovarian cancer is scarce. This study aimed to evaluate chemotherapy adherence, reasons for chemotherapy modifications, and associations with overall survival.

**Methods:** Advanced-stage epithelial ovarian cancer patients diagnosed between January 2015 and December 2021 were identified from the Netherlands Cancer Registry. Patients who underwent cytoreductive surgery combined with platinum- and taxane-based chemotherapy were included. Patients were categorized into two groups: adherent (patients without modifications) and non-adherent (patients with modifications: dose reduction, chemotherapy interruption, and/or reduction in chemotherapy cycles). Reasons for modifications were assessed. Kaplan-Meier survival curves and Cox proportional hazards models were used to analyze overall survival.

**Results:** Among the cohort (N = 3,687), 54% of patients underwent chemotherapy modifications. Dose reduction (38%) was the most common, followed by interruption (24%) and reduction in chemotherapy cycles (9%). Non-adherence was associated with poorer performance scores, higher comorbidity indices, and undergoing primary cytoreductive surgery. Neurotoxicity and hematologic toxicity were the primary reasons for modifications in platinum (33% and 37%) and taxane (47% and 35%) agents. No association with survival was found for dose reduction and interruption. However, reduction in chemotherapy cycles was associated with lower 5-year overall survival (32% (95% CI 26%-38%) vs. 36% (95% CI 34%-38%)), remaining significant after multivariable adjustment (hazard ratio 1.36; 95% CI 1.17-1.59).

**Conclusion:** A significant proportion of Dutch advanced-stage epithelial ovarian cancer patients undergo chemotherapy modifications. No impact on overall survival was found for dose reduction or chemotherapy interruption, warranting prospective studies. Reduction in chemotherapy cycles was negatively associated with overall survival, possibly reflecting underlying treatment ineffectiveness.

**Key messages:** *What is already known on this topic:* Guideline-recommended chemotherapy for advanced epithelial ovarian cancer is often difficult to deliver in routine practice, and real-world data on adherence and its impact on survival are limited.

*What this study adds:* In this nationwide retrospective cohort, over half of patients experienced chemotherapy modifications; dose reductions and interruptions were not associated with poorer overall survival, whereas a reduction in the number of cycles showed an association with worse outcomes, although this may partly reflect underlying disease severity or treatment response.

*How this study might affect research, practice or policy:* Our findings suggest that standard dosing and treatment duration of six cycles may not always be necessary, emphasizing the need to tailor treatment plans to optimize both efficacy and tolerability in advanced-stage epithelial ovarian cancer patients

## Introduction

Chemotherapy is one of the cornerstones in the treatment of primary epithelial ovarian cancer [1]. In advanced-stage disease, the European guidelines state primary cytoreductive surgery (PCS) followed by six cycles of chemotherapy as the gold standard of treatment [1, 2]. First-line chemotherapy consists of paclitaxel (175 mg/m^2^) and carboplatin (AUC 5–6) administered every three weeks for a total of six cycles[1, 2]. Alternatively, when complete cytoreduction is not deemed feasible (e.g., due to spread of disease or the patient’s general condition), three cycles of neoadjuvant chemotherapy, followed by interval cytoreductive surgery (NACT-ICS) and three adjuvant chemotherapy cycles, are recommended [1, 2].

Despite clear guidelines, variation in chemotherapy administration has been reported for epithelial ovarian cancer [3]. For instance, a French multicenter study disclosed that only 44% of patients underwent guideline-recommended chemotherapy [4]. In an Australian nationwide study, Jordan et al. reported that 68% of patients received platinum-based chemotherapy combined with taxanes [5]. Furthermore, it was reported that only 50% of patients completed the recommended six cycles without modifications, and 68% of patients aged over 70 did not undergo the standard chemotherapy regimen [5]. Most studies, except for Jordan et al., are not nationwide investigations or are limited by small sample sizes [3–7]. The lack of nationwide studies may imply the potential existence of disparities in clinical practices, that remain inadequately explored in the current literature.

Therefore, the aim of this nationwide study was to assess advanced-stage epithelial ovarian cancer patients’ adherence to first-line chemotherapy, as recommended by the European guidelines, in the Netherlands. Furthermore, reasons for chemotherapy modifications (i.e., dose reduction, chemotherapy interruption, or reduction in number of chemotherapy cycles) and their impact on patients’ overall survival were assessed.

## Methods

### Data collection

In this retrospective cohort study, patients with peritoneal, ovarian and fallopian tube cancers (i.e., International Classification of Disease for Oncology [ICD-O-3] codes C48.1, C48.2, C56.9, and C57.0), diagnosed between January 1, 2015 and December 31, 2021 were identified from the Netherlands Cancer Registry (NCR). That is a population-based registry that receives weekly notifications of all cytologically and histologically confirmed malignancies in the Netherlands through an automated nationwide pathology archive (PALGA). Dedicated data managers extract data on patient, tumor and treatment characteristics from medical records. To obtain recent information on vital status and date of death, the NCR is annually linked to the Personal Records Database.

### Study population

Patients primarily diagnosed with advanced-stage epithelial ovarian cancer (i.e. International Federation of Gynecology and Obstetrics (FIGO) stages IIB–IV) were identified. Solely advanced-stage patients who underwent cytoreductive surgery combined with at least one cycle of platinum- and taxane-based chemotherapy were included. Patients who underwent another form of treatment were excluded. In addition, patients with missing data on chemotherapy regimens or modifications were also excluded from this study.

### Definitions

Chemotherapy modifications comprised chemotherapy dose reduction, chemotherapy interruption, or reduction in the number of chemotherapy cycles. Dose adjustments based on the decreased estimated glomerular filtration rates or patients’ weight changes after ascites drainage were not considered dose reductions. Overall survival was defined as the time between the date of diagnosis and the date of death, or last follow-up for patients who were still alive (censoring date: January 31, 2024). The World Health Organization (WHO) performance score was used as a performance status. Residual disease was defined as the maximum diameter of the largest tumor nodule remaining after cytoreductive surgery, classified as no macroscopic (complete cytoreduction), ≤1 cm (optimal cytoreduction), or >1 cm (incomplete cytoreduction) residual disease.

### Statistical analysis

Patient, tumor, and treatment characteristics were summarized using descriptive statistics. Patients who had chemotherapy modifications, along with the reasons for these modifications, were identified. Patients were divided into adherent (patients without chemotherapy modifications) and non-adherent (patients with chemotherapy modifications) groups. To compare the two groups, Pearson χ² or Fisher’s exact tests were used for categorical variables, and two-sample Wilcoxon rank-sum tests were used for continuous variables. To assess whether chemotherapy modifications affect overall survival, survival analyses were conducted using Kaplan-Meier survival curves and multivariable Cox proportional hazards models. Survival was adjusted for performance status, histologic subtype, tumor grade, FIGO stage, treatment approach (i.e., primary cytoreductive surgery or neoadjuvant chemotherapy followed by interval cytoreductive surgery), residual disease, type of chemotherapy modification, use of hyperthermic intraperitoneal chemotherapy (HIPEC), and use of poly(ADP-ribose) polymerase (PARP) inhibitors. Furthermore, a subgroup analysis was conducted to compare survival between patients who received five chemotherapy cycles and those who received six cycles. This analysis evaluated whether omitting the final chemotherapy cycle impacts patient survival, as this is often considered in clinical practice for several reasons. Two-sided *p*-values of less than 0.05 were considered statistically significant. All statistical analyses were performed using STATA/SE, version 17.1 (StataCorp, College Station, TX, USA).

### Ethical approval

Ethical approval for this study was obtained from the NCR’s Privacy Review Board [K23.306].

## Results

A total of 9,082 patients were diagnosed with primary epithelial ovarian cancer between January 1, 2015 and December 31, 2021. Of these, 3,687 (41%) had advanced-stage disease and underwent cytoreductive surgery combined with platinum- and taxane-based chemotherapy. Among them, 1,713 patients (46%) underwent treatment without chemotherapy modifications (adherent group), while 1,974 patients (54%) underwent treatment with chemotherapy modifications (non-adherent group), see **Figure 1**.

**Figure 1.**
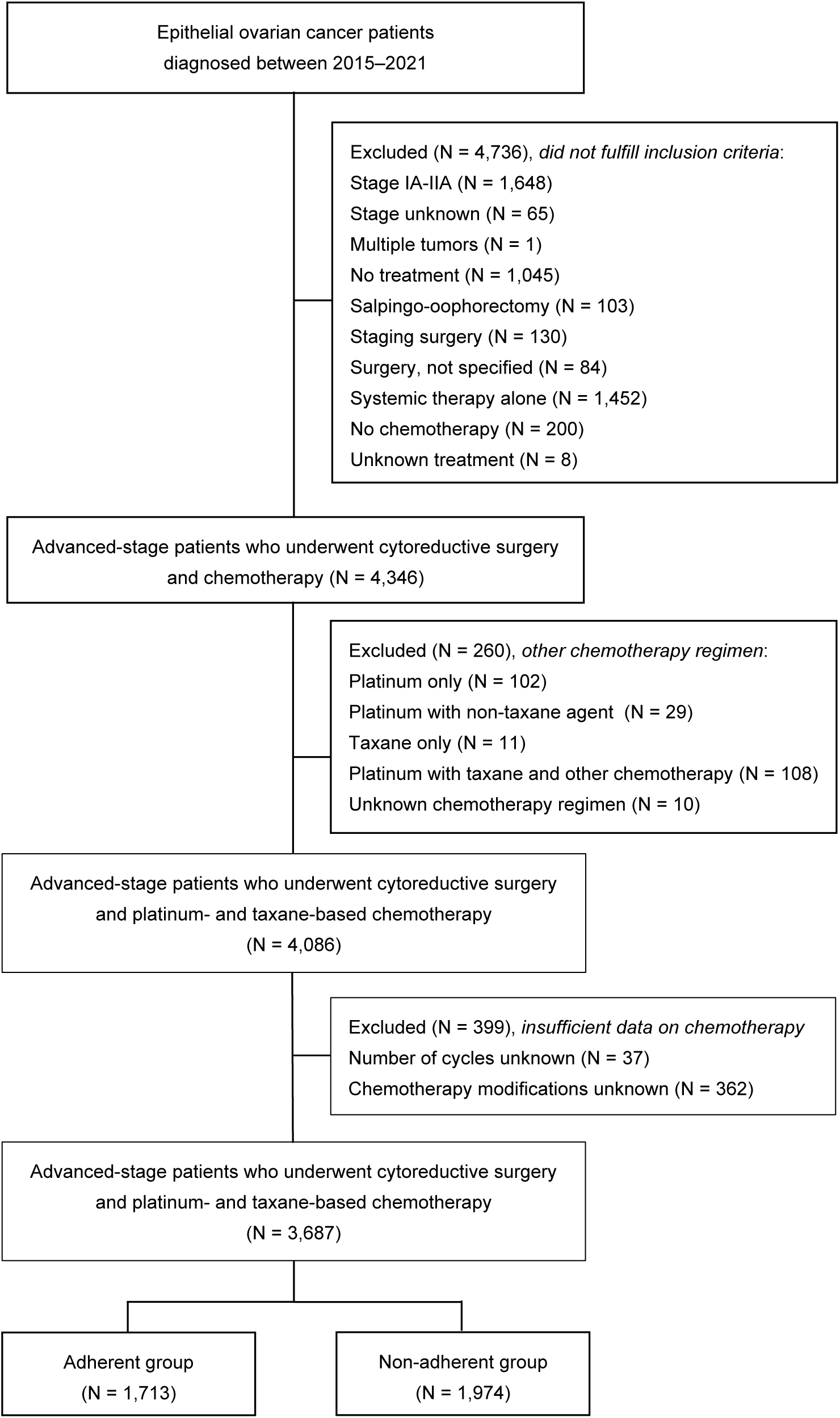
Flowchart of the study population.

**Table 1** presents the patient, tumor, and treatment characteristics of the study cohort. The median age at diagnosis was 66 years (interquartile range [IQR], 59–72), and the most common WHO performance score was 0 (45%). Serous carcinoma was the most prevalent histologic subtype (84%). The majority of patients underwent neoadjuvant chemotherapy followed by interval cytoreductive surgery (72%). Compared with patients who did not undergo chemotherapy modifications, those with modifications generally had more comorbidities, worse performance scores, and were more likely to undergo primary cytoreductive surgery.

**Table 1.**
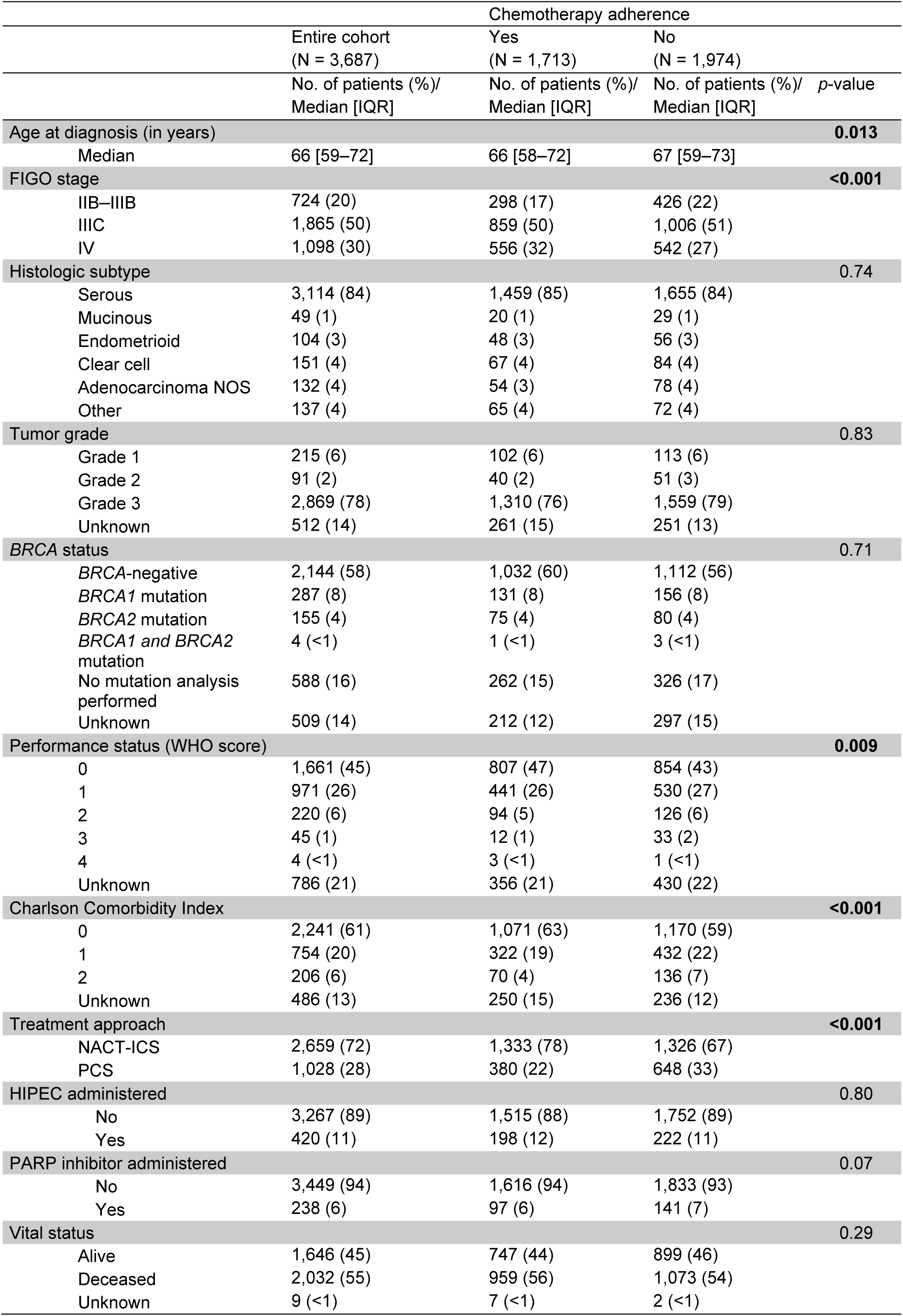
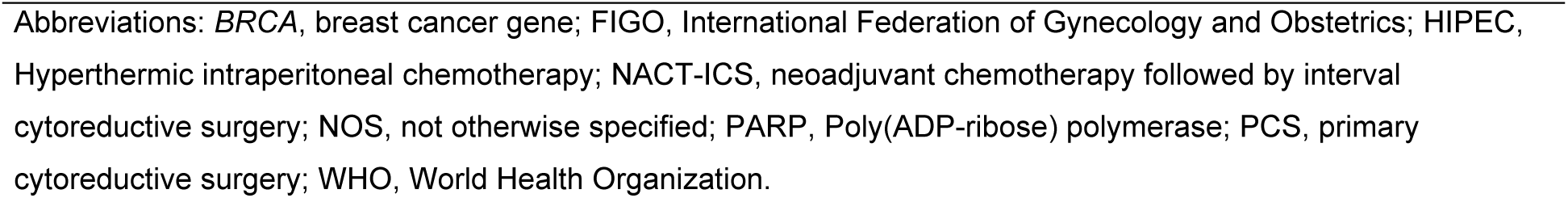
Patient, tumor, and treatment characteristics of the entire study cohort (N = 3,687), also stratified by chemotherapy adherence.

|  | Entire cohort<br>(N = 3,687) | Chemotherapy adherence |  | p-value |
| --- | --- | --- | --- | --- |
|  |  | Yes<br>(N = 1,713) | No<br>(N = 1,974) |  |
|  | No. of patients (%)/<br>Median [IQR] | No. of patients (%)/<br>Median [IQR] | No. of patients (%)/<br>Median [IQR] |  |
| Age at diagnosis (in years) |  |  |  | **0.013** |
| Median | 66 [59–72] | 66 [58–72] | 67 [59–73] |  |
| FIGO stage |  |  |  | **<0.001** |
| IIB–IIIB | 724 (20) | 298 (17) | 426 (22) |  |
| IIIC | 1,865 (50) | 859 (50) | 1,006 (51) |  |
| IV | 1,098 (30) | 556 (32) | 542 (27) |  |
| Histologic subtype |  |  |  | 0.74 |
| Serous | 3,114 (84) | 1,459 (85) | 1,655 (84) |  |
| Mucinous | 49 (1) | 20 (1) | 29 (1) |  |
| Endometrioid | 104 (3) | 48 (3) | 56 (3) |  |
| Clear cell | 151 (4) | 67 (4) | 84 (4) |  |
| Adenocarcinoma NOS | 132 (4) | 54 (3) | 78 (4) |  |
| Other | 137 (4) | 65 (4) | 72 (4) |  |
| Tumor grade |  |  |  | 0.83 |
| Grade 1 | 215 (6) | 102 (6) | 113 (6) |  |
| Grade 2 | 91 (2) | 40 (2) | 51 (3) |  |
| Grade 3 | 2,869 (78) | 1,310 (76) | 1,559 (79) |  |
| Unknown | 512 (14) | 261 (15) | 251 (13) |  |
| BRCA status |  |  |  | 0.71 |
| BRCA-negative | 2,144 (58) | 1,032 (60) | 1,112 (56) |  |
| BRCA1 mutation | 287 (8) | 131 (8) | 156 (8) |  |
| BRCA2 mutation | 155 (4) | 75 (4) | 80 (4) |  |
| BRCA1 and BRCA2<br>mutation | 4 (<1) | 1 (<1) | 3 (<1) |  |
| No mutation analysis<br>performed | 588 (16) | 262 (15) | 326 (17) |  |
| Unknown | 509 (14) | 212 (12) | 297 (15) |  |
| Performance status (WHO score) |  |  |  | **0.009** |
| 0 | 1,661 (45) | 807 (47) | 854 (43) |  |
| 1 | 971 (26) | 441 (26) | 530 (27) |  |
| 2 | 220 (6) | 94 (5) | 126 (6) |  |
| 3 | 45 (1) | 12 (1) | 33 (2) |  |
| 4 | 4 (<1) | 3 (<1) | 1 (<1) |  |
| Unknown | 786 (21) | 356 (21) | 430 (22) |  |
| Charlson Comorbidity Index |  |  |  | **<0.001** |
| 0 | 2,241 (61) | 1,071 (63) | 1,170 (59) |  |
| 1 | 754 (20) | 322 (19) | 432 (22) |  |
| 2 | 206 (6) | 70 (4) | 136 (7) |  |
| Unknown | 486 (13) | 250 (15) | 236 (12) |  |
| Treatment approach |  |  |  | **<0.001** |
| NACT-ICS | 2,659 (72) | 1,333 (78) | 1,326 (67) |  |
| PCS | 1,028 (28) | 380 (22) | 648 (33) |  |
| HIPEC administered |  |  |  | 0.80 |
| No | 3,267 (89) | 1,515 (88) | 1,752 (89) |  |
| Yes | 420 (11) | 198 (12) | 222 (11) |  |
| PARP inhibitor administered |  |  |  | 0.07 |
| No | 3,449 (94) | 1,616 (94) | 1,833 (93) |  |
| Yes | 238 (6) | 97 (6) | 141 (7) |  |
| Vital status |  |  |  | 0.29 |
| Alive | 1,646 (45) | 747 (44) | 899 (46) |  |
| Deceased | 2,032 (55) | 959 (56) | 1,073 (54) |  |
| Unknown | 9 (<1) | 7 (<1) | 2 (<1) |  |

Abbreviations: *BRCA*, breast cancer gene; FIGO, International Federation of Gynecology and Obstetrics; HIPEC, Hyperthermic intraperitoneal chemotherapy; NACT-ICS, neoadjuvant chemotherapy followed by interval cytoreductive surgery; NOS, not otherwise specified; PARP, Poly(ADP-ribose) polymerase; PCS, primary cytoreductive surgery; WHO, World Health Organization.

In the entire study cohort, patients were more likely to receive the guideline-recommended six cycles of platinum than six cycles of taxane. **Supplementary Figures 1–3** demonstrate the number of chemotherapy cycles administered after primary cytoreductive surgery, and before and after interval cytoreductive surgery, respectively. In the primary cytoreductive surgery setting (N = 1,028, 28%), six cycles of adjuvant platinum were administered to 89% of patients, while six cycles of taxane were administered to 81% of the patients (p<0.001) (**Supplementary Figure 1**).

In the interval cytoreductive surgery setting (N = 2,659, 72%), the proportion of patients undergoing the recommended three cycles of neoadjuvant chemotherapy was similar for both agents (platinum 79% vs. taxane 78%) (**Supplementary Figure 2**). However, the proportion of patients who underwent the recommended three cycles of adjuvant chemotherapy after neoadjuvant chemotherapy followed by interval cytoreductive surgery was higher for platinum (83%) than taxane (78%) (p<0.001) (**Supplementary Figure 3**).

**Table 2** demonstrates the types of chemotherapy modifications across the entire cohort, as well as stratified by chemotherapy agent. Dose reduction was the most commonly reported chemotherapy modification (38%), followed by chemotherapy interruption (24%), and reduction in the number of chemotherapy cycles (9%). Multiple types of chemotherapy modifications were reported for 624 patients (17%). Dose reduction (31% vs. 36%, p<0.001) and reduction in the number of chemotherapy cycles (4% vs. 8%, p<0.001) were less frequently reported for platinum agents compared to taxane agents. **Supplementary Table 1** further stratifies the chemotherapy modifications by chemotherapy timing (neoadjuvant vs. adjuvant).

**Table 2.**
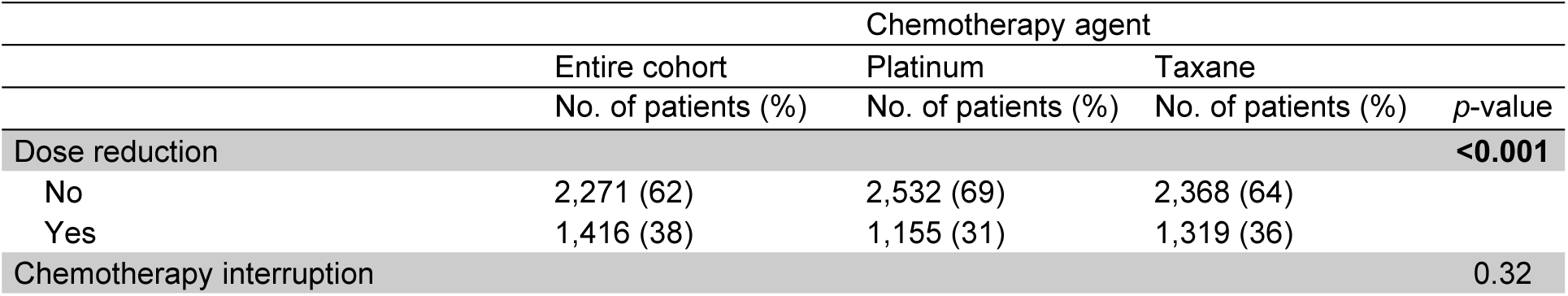
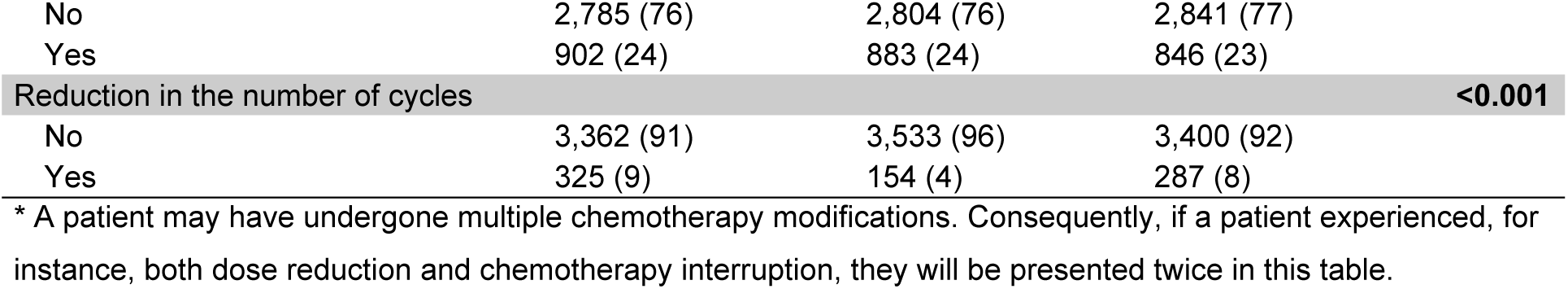
The types of chemotherapy modifications across the entire cohort (N = 3,687), also stratified by chemotherapy agent.

The reasons for chemotherapy modifications in patients are listed in **Figure 2**. Neurotoxicity was the predominant reason involving taxane agents (47%) and the second most common for platinum agents (33%). Conversely, hematologic toxicity was the most common reason for modifications involving platinum agents (37%) and the second most frequent for taxane agents (35%). The reasons for each modification type are detailed in **Supplementary Figures 4–6**. Neurotoxicity was the most frequently reported reason for dose reduction (51% for platinum vs. 63% for taxane) and for reduction in the number of chemotherapy cycles in taxane agents. In contrast, reasons other than those listed were most commonly reported for reduction in chemotherapy cycles for platinum agents (39%). Hematologic toxicity was the predominant reason for chemotherapy interruption in both platinum (64%) and taxane (63%) agents.

**Figure 2.**
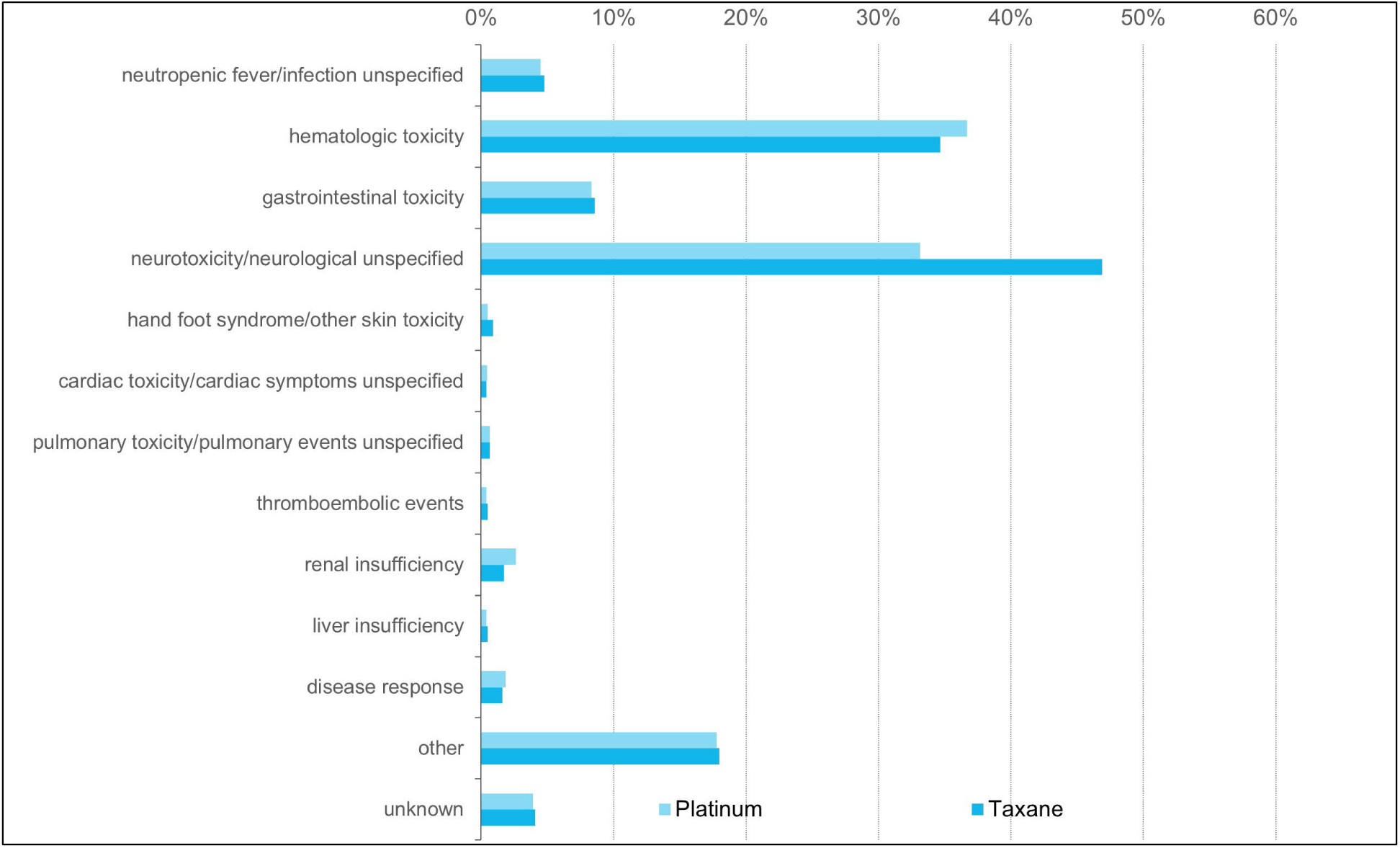
The reasons for chemotherapy modifications (i.e., dose reduction, chemotherapy interruption, or reduction in the number of cycles combined) of the entire cohort. *Patients could have had chemotherapy modifications due to more than one reason. †The other category comprises reasons for chemotherapy modifications other than one of the listed reasons.

Kaplan-Meier survival curves are presented in **Figure 3**. The 5-year survival rate of the entire cohort was 36% (95% confidence interval (CI) 34%–38%). The 5-year survival rates of patients with and without dose reduction were similar (36% (95–CI 33%–39%) vs. 35% (95%–CI 33%–38%)). Similarly, no notable difference in the 5-year survival was found between patients with and without chemotherapy interruption (36% (95–CI 32%–39%) vs. 36% (95%–CI 33%–38%)). However, patients with reduction in the number of chemotherapy cycles had significantly lower 5-year survival rates than those without (32% (95–CI 26%–38%) vs. (]36% (95%–CI 34%–38%)), even after multivariable adjustment (hazard ratio (HR) 1.36; 95%–CI 1.17–1.59).

**Figure 3.**
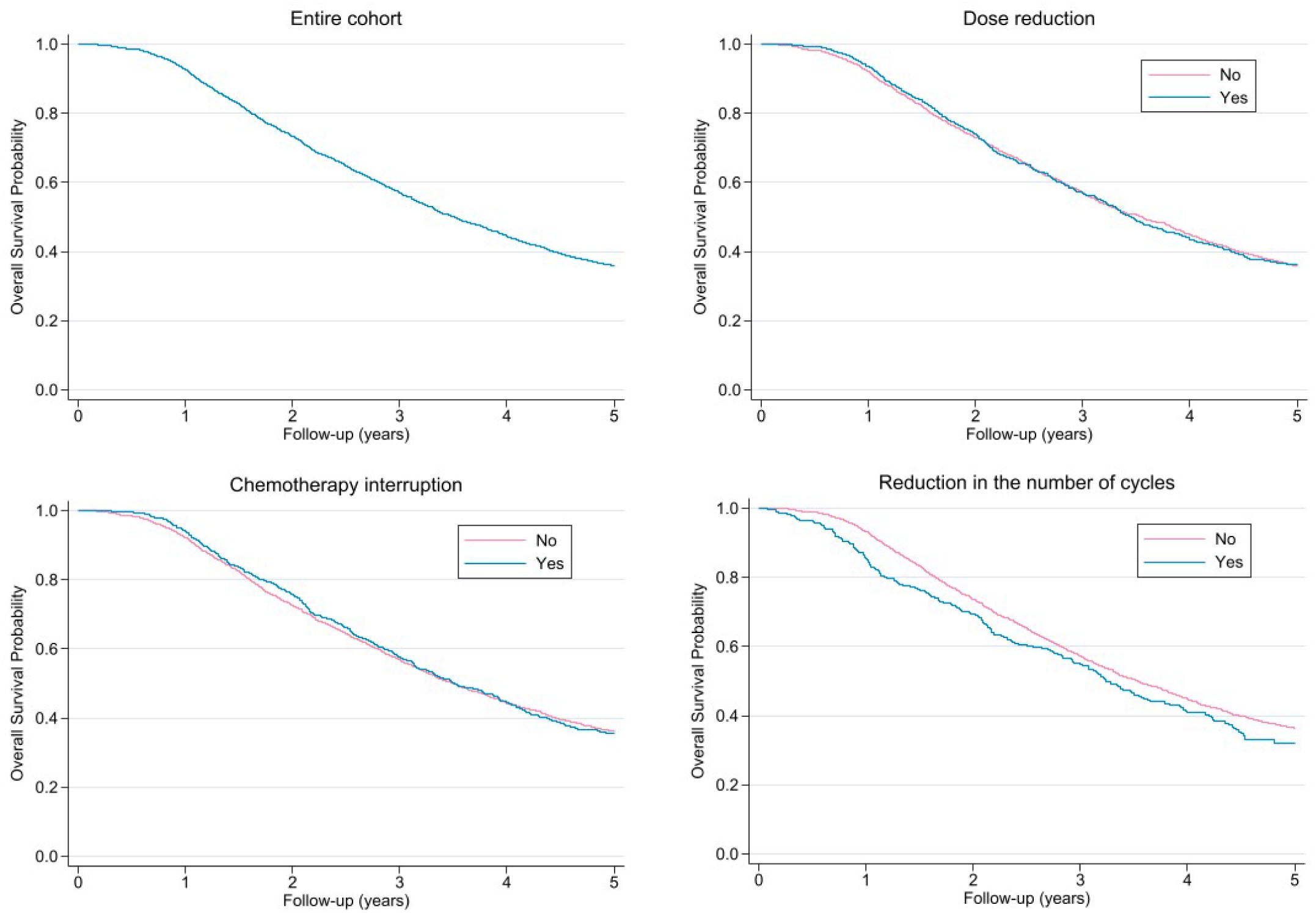
Kaplan-Meier survival curves for the entire cohort and for patients with or without dose reductions, chemotherapy interruptions, or reductions in the number of chemotherapy cycles. *Hazard ratios (HR) from multivariable Cox proportional hazards models: dose reduction (HR 1.09, 95%–CI: 0.98–1.19); chemotherapy interruption (HR 0.98, 95%–CI: 0.88–1.09); reduction in the number of chemotherapy cycles (HR 1.36, 95%–CI: 1.17–1.59).

A subgroup analysis was conducted to determine whether omitting the final chemotherapy cycle affects patient survival, as reducing the number of cycles is often considered in clinical practice to minimize toxicity or manage side effects. The Kaplan-Meier survival curves of patients who received five vs. six chemotherapy cycles in the primary cytoreductive surgery and neoadjuvant chemotherapy followed by interval cytoreductive surgery setting are presented in **Supplementary Figure 7.** No notable survival difference was observed between patients who underwent five or six chemotherapy cycles in the primary cytoreductive surgery setting (HR 1.53 95%–CI 0.84–2.76). Similarly, no significant difference in survival was found between patients who underwent five or six chemotherapy cycles in the neoadjuvant chemotherapy followed by interval cytoreductive surgery setting (HR 1.25 95%–CI 0.88–1.76).

## Discussion

### Summary of Main Results

This nationwide cohort study provides significant insights into the chemotherapy adherence of advanced-stage epithelial ovarian cancer patients in the Netherlands. Our findings confirm that variability in the administration of chemotherapy is common among these patients, with more than half (54%) experiencing adjustments to their treatment regimens. While dose reduction and chemotherapy interruption (or delay) do not appear to negatively impact overall survival, reduction in the number of chemotherapy cycles, specifically more than one reduced cycle, may be associated with decreased overall survival.

### Results in the Context of Published Literature

Dose reduction emerged as the most common chemotherapy modification among patients, often attributed to neurotoxicity. Surprisingly, despite receiving lower doses than initially intended, these patients did not experience worse survival outcomes. This finding is consistent with the results of Lee et al. (N = 102) and Nagel et al. (N = 175), who also found no negative impact of dose reductions on overall survival in epithelial ovarian cancer patients [8]. Particularly, Lee et al. demonstrated that even dose reductions of ≥60% did not adversely affect overall or progression-free survival in advanced-stage disease [8]. These findings prompt further investigation into whether the initial dosing regimens are higher than necessary, given that dose reduction does not appear to compromise overall survival and may be associated with avoidable adverse events.

While reports on the impact of chemotherapy interruption or delay on epithelial ovarian cancer survival have been inconsistent [9–12], our results suggest that it is feasible to manage adverse effects without comprising the efficacy of treatment. Consistently, Starbuck et al. (N = 505) demonstrated similar survival between patients without chemotherapy delays and patients with a prolonged treatment duration of less than four weeks. However, their study showed that survival probabilities decreased when the total delay in treatment exceeded six weeks [12]. In addition, Searle et al. (N = 205) reported that a chemotherapy interruption of more than ten weeks was associated with poorer survival [10]. The lack of association with survival in our cohort may be attributed to better management of treatment intervals in the Netherlands.

The number of patients undergoing a reduction in chemotherapy cycles was relatively low (4% for platinum and 8% for taxane). Among all chemotherapy modifications, only a decrease in the number of chemotherapy cycles was associated with poorer overall survival. The proportion of patients with disease progression or lack of treatment response was higher among those with reduced chemotherapy cycles compared to those with dose reductions or chemotherapy interruptions. Therefore, the poorer survival outcomes observed in the patients with reduced chemotherapy cycles may be attributed to the response of the treatment rather than the reduced number of cycles alone. However, it is important to note that, when adjusting for performance status, histologic subtype, tumor grade, FIGO stage, treatment approach, residual disease, and type of chemotherapy modification, no significant difference in survival was found between patients receiving five or six cycles. This suggests that a reduction of more than one chemotherapy cycle may be necessary to negatively impact survival outcomes.

### Strengths and Weaknesses

Our study is strengthened by its large sample size and nationwide population-based design, offering a representative overview of chemotherapy adherence among epithelial ovarian cancer patients in the Netherlands. Moreover, our study provides comprehensive insights into chemotherapy modifications, serving as one of the first nationwide studies to explore the reasons behind these modifications and their impact on survival. Nonetheless, limitations of our study include the lack of data on the extent of dose reductions and detailed information regarding the median duration of chemotherapy interruption. Furthermore, due to the retrospective observational nature of the data, the ability to draw definitive conclusions is limited.

### Implications for Practice and Future Research

Our findings suggest that standard dosing and treatment duration of six cycles may not always be necessary, emphasizing the need to tailor treatment plans to optimize both efficacy and tolerability in advanced-stage epithelial ovarian cancer patients. Clinical recommendations are often derived from studies that may not fully represent the diverse patient population seen in real-world clinical practice. If a substantial proportion of patients deviates from standard treatment regimens, recommendations based solely on those regimens may have limited practicality. Bridging the gap between clinical research and real-world practice using population-based research is crucial for improving chemotherapy adherence and outcomes. This may involve inclusive study designs, guidelines accommodating patient variability, and supportive interventions addressing adherence barriers. Personalized treatment approaches, considering patient factors while adhering closely to guidelines, are essential. Additionally, strategies to manage chemotherapy-related toxicities could mitigate the need for treatment modifications.

## Conclusions

In conclusion, this nationwide study highlights significant variability in chemotherapy adherence among advanced-stage epithelial ovarian cancer patients in the Netherlands, with a substantial proportion of patients undergoing chemotherapy modifications. Chemotherapy dose reduction and interruption were not negatively associated with overall survival, suggesting the need for further prospective studies to explore potential adjustments to dosing schedules. Conversely, reduction in the number of chemotherapy cycles, more than one reduced cycle, was associated with worse overall survival, possibly due an inadequate response to chemotherapy.

## Data Availability

All data produced in the present study are available upon reasonable request to the authors

## Acknowledgements

The authors thank the Netherlands Cancer Registry for collecting and providing the data used in this study.

## CRediT author statement

S.A. Said: Conceptualization, Methodology, Formal analysis, Investigation, Visualization, Writing – Original Draft, Writing – Review & Editing

H.H.B Wenzel: Conceptualization, Methodology, Formal analysis, Investigation, Visualization, Writing – Review & Editing

A.M. van Altena: Conceptualization, Methodology, Supervision, Visualization, Writing – Review & Editing

J.E.W. Walraven: Conceptualization, Visualization, Writing – Review & Editing

J. IntHout: Conceptualization, Methodology, Supervision, Visualization, Writing – Review & Editing

J. A. de Hullu: Conceptualization, Methodology, Supervision, Visualization, Writing – Review & Editing

M.A. van der Aa: Conceptualization, Methodology, Supervision, Visualization, Writing – Review & Editing

## Author disclosure statement

No disclosures are reported for all authors.

## Funding

This work was supported by Dutch Cancer Society [IKNL2014-6838].

## Supplementary

**Supplementary table 1.** Types of chemotherapy modifications reported for patients who underwent neoadjuvant (N = 2,659) and/or adjuvant (N=3,545) chemotherapy, stratified by chemotherapy agent.

|  | Neoadjuvant |  |  |  | Adjuvant |  |  |  |
| --- | --- | --- | --- | --- | --- | --- | --- | --- |
|  | Total | Platinum | Taxane |  | Total | Platinum | Taxane |  |
|  | No. of patients (%) | No. of patients (%) | No. of patients (%) | p-value | No. of patients (%) | No. of patients (%) | No. of patients (%) | p-value |
| Dose reduction |  |  |  | **0.002** |  |  |  | **<0.001** |
| No | 1,723 (65) | 1,891 (71) | 1,783 (67) |  | 2,170 (61) | 2,424 (68) | 2,265 (64) |  |
| Yes | 936 (35) | 768 (29) | 871 (33) |  | 1,375 (39) | 1,121 (32) | 1,280 (36) |  |
| Chemotherapy interruption |  |  |  | 0.49 |  |  |  | 0.31 |
| No | 2,036 (77) | 2,047 (77) | 2,069 (78) |  | 2,670 (75) | 2,689 (76) | 2,726 (77) |  |
| Yes | 623 (23) | 612 (23) | 590 (22) |  | 875 (25) | 856 (24) | 819 (23) |  |
| Reduction in the number of cycles |  |  |  | **<0.001** |  |  |  | **<0.001** |
| No | 2,478 (93) | 2,578 (97) | 2,499 (94) |  | 3,227 (91) | 3,393 (96) | 3,265 (92) |  |
| Yes | 181 (7) | 81 (3) | 160 (6) |  | 318 (9) | 152 (4) | 280 (8) |  |

**Supplementary Figure 1.**
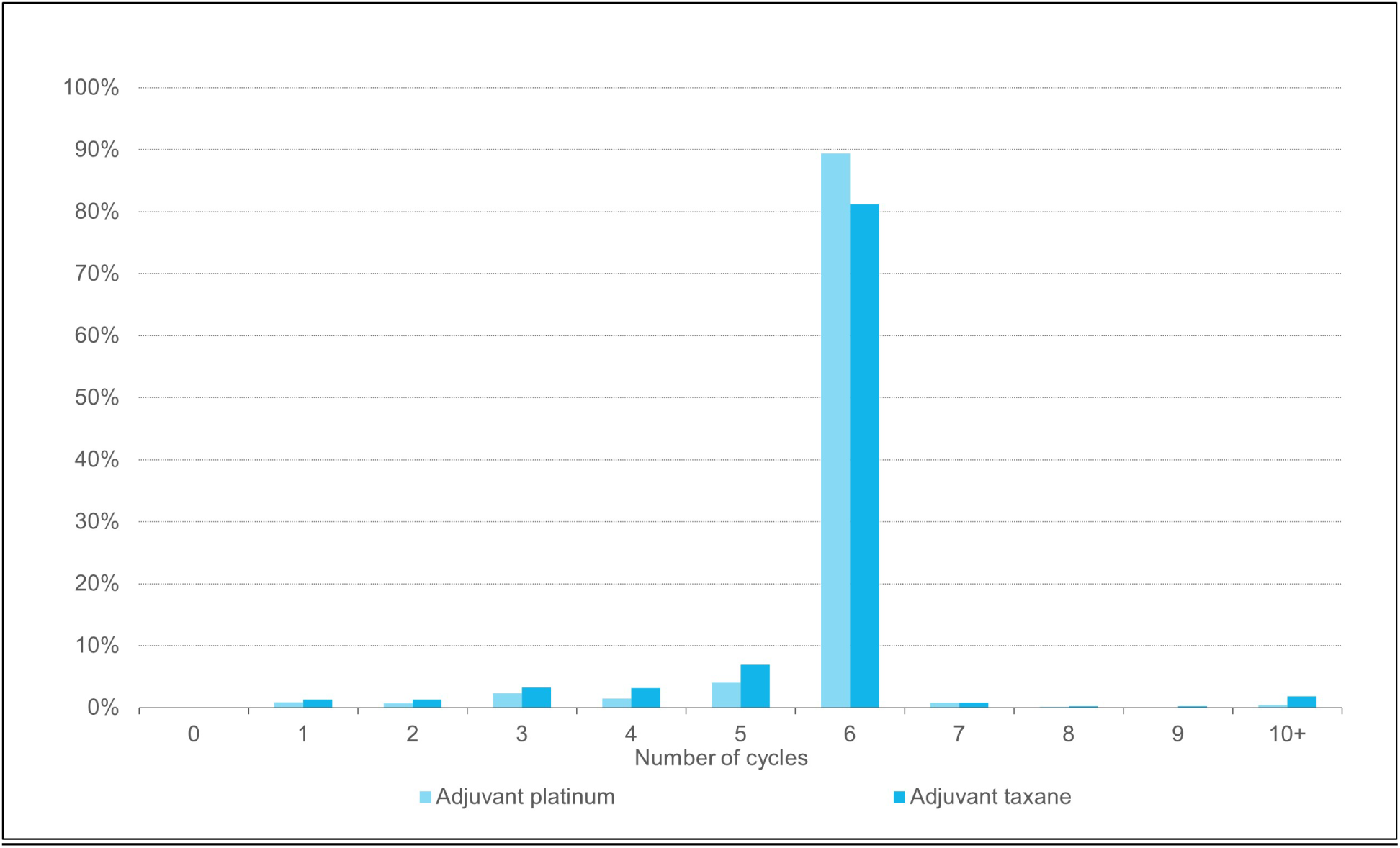
The proportion of patients by the number of adjuvant chemotherapy cycles in the primary cytoreductive setting (N = 1,028).

**Supplementary Figure 2.**
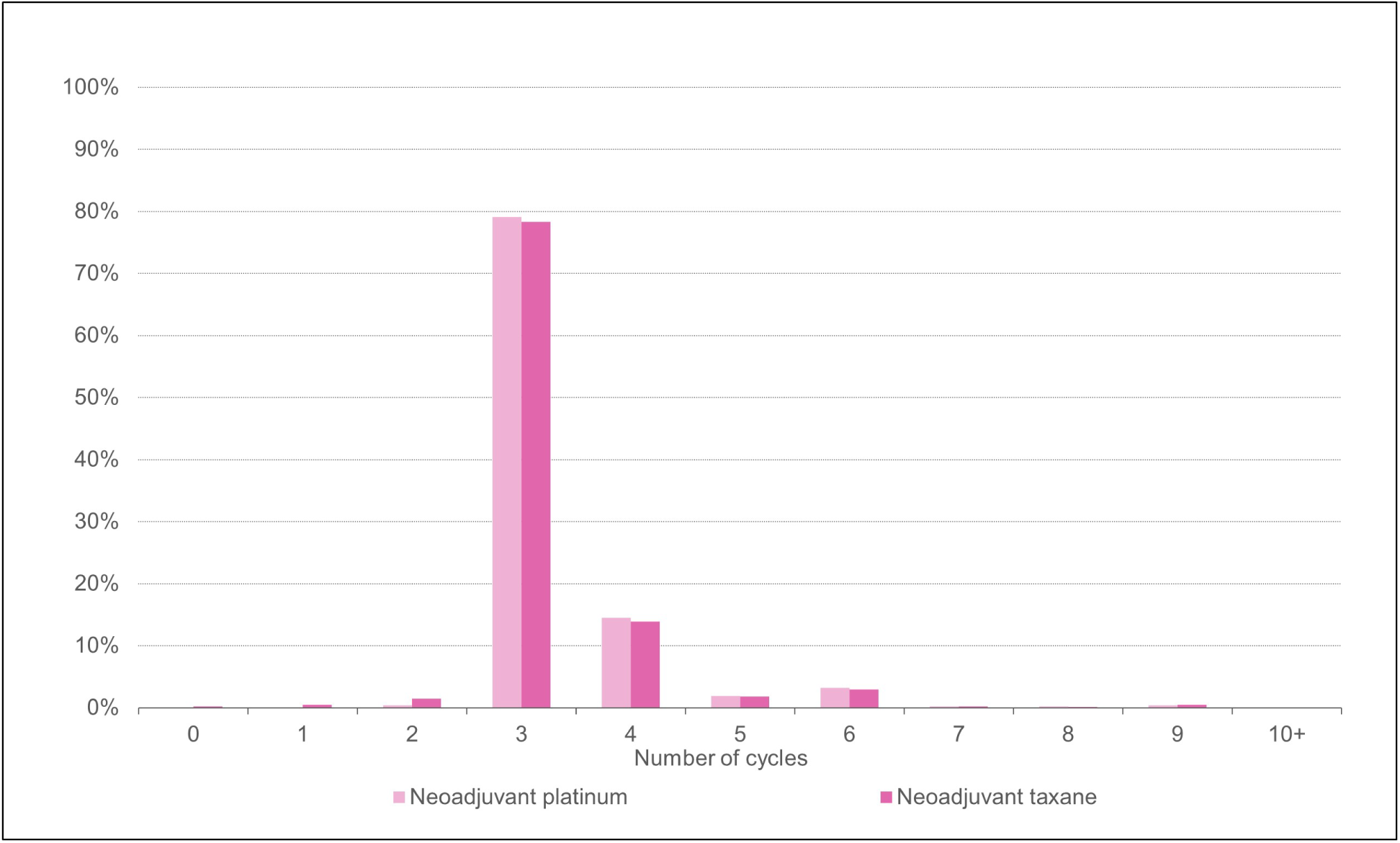
The proportion of patients by the number of neoadjuvant chemotherapy cycles in the interval cytoreductive setting (N = 2,659).

**Supplementary Figure 3.**
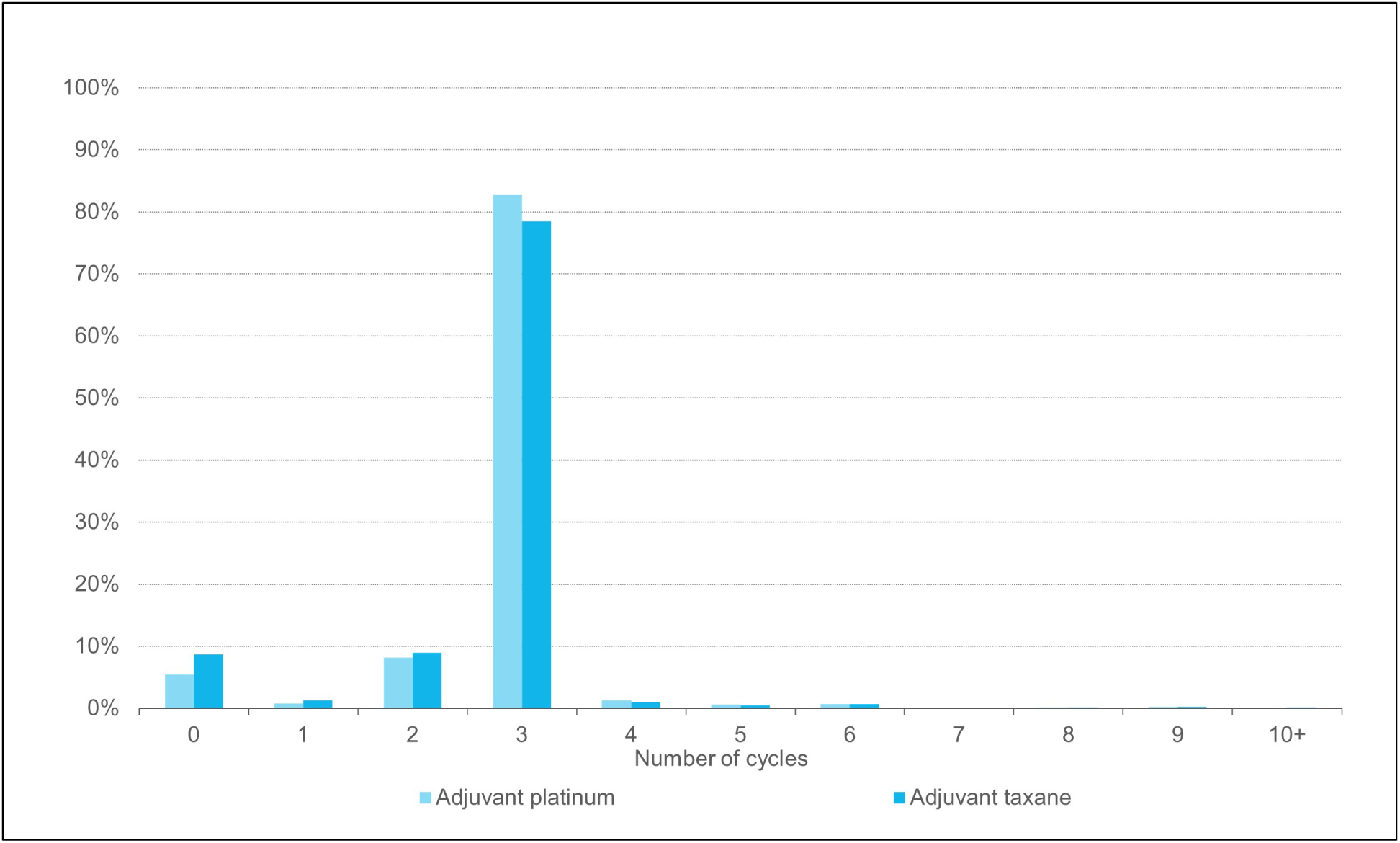
The proportion of patients by the number of adjuvant chemotherapy cycles in the interval cytoreductive setting (N = 2,659).

**Supplementary Figure 4.**
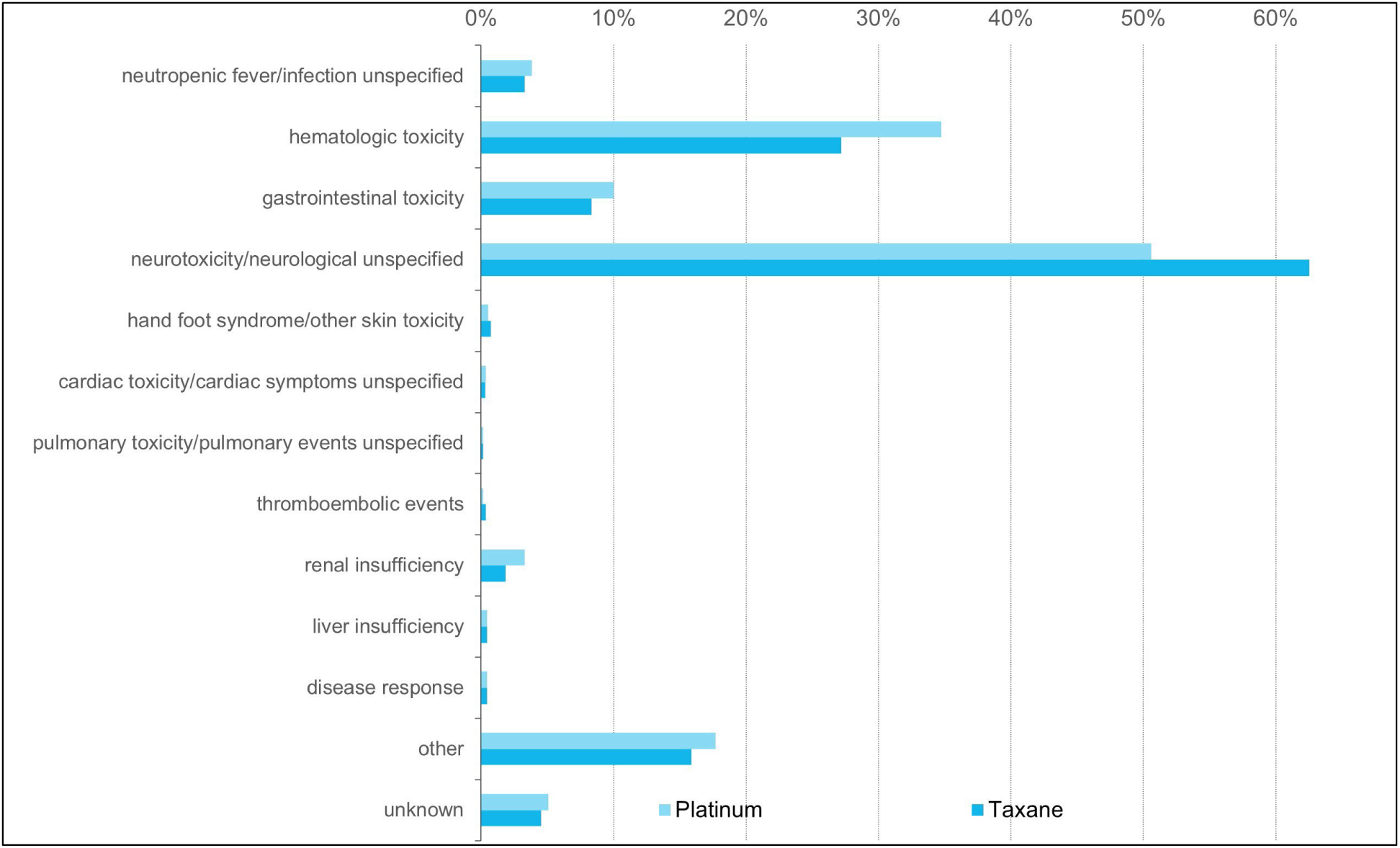
Reasons for dose reductions in the entire cohort.

**Supplementary Figure 5.**
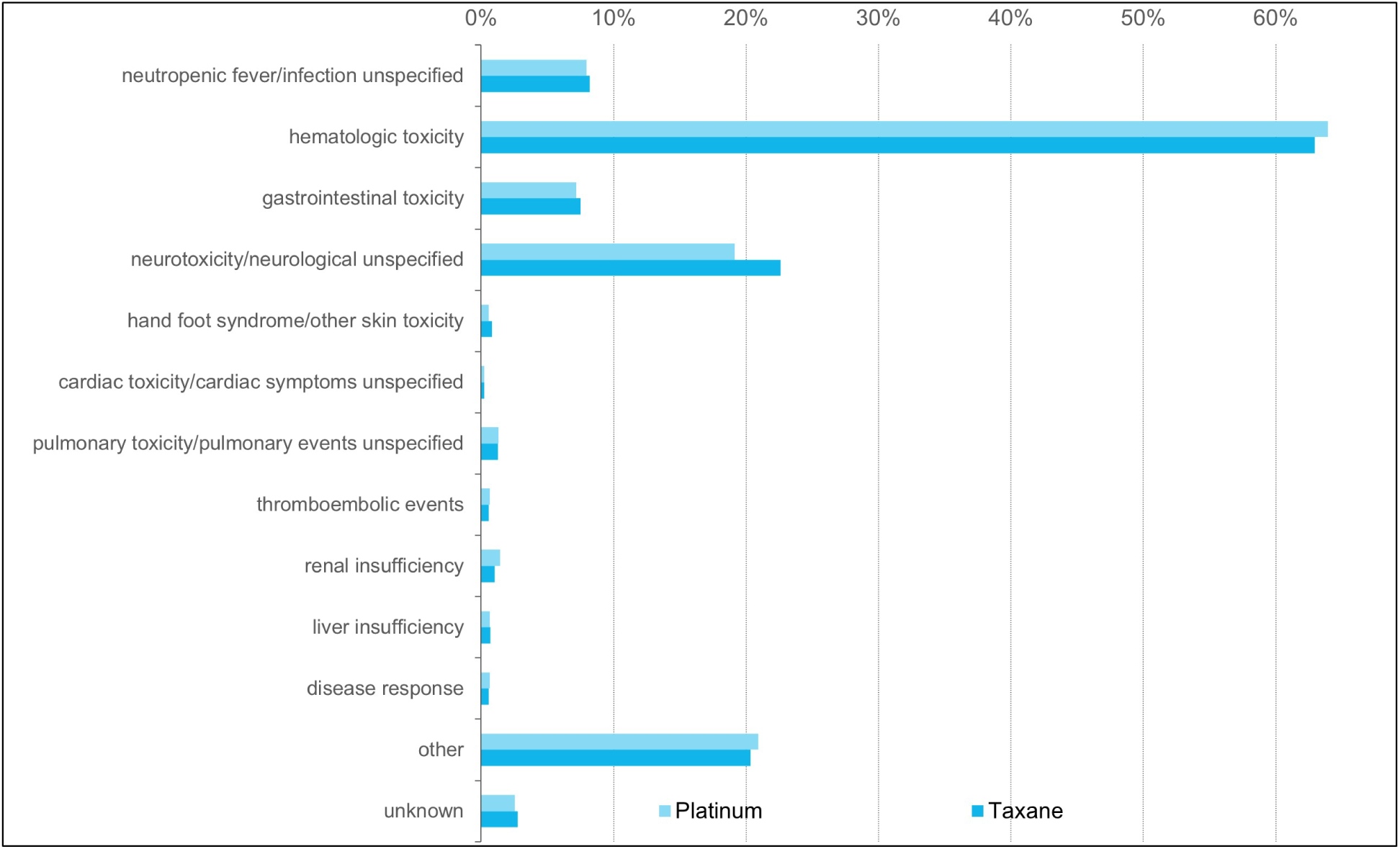
Reasons for chemotherapy interruptions in the entire cohort.

**Supplementary Figure 6.**
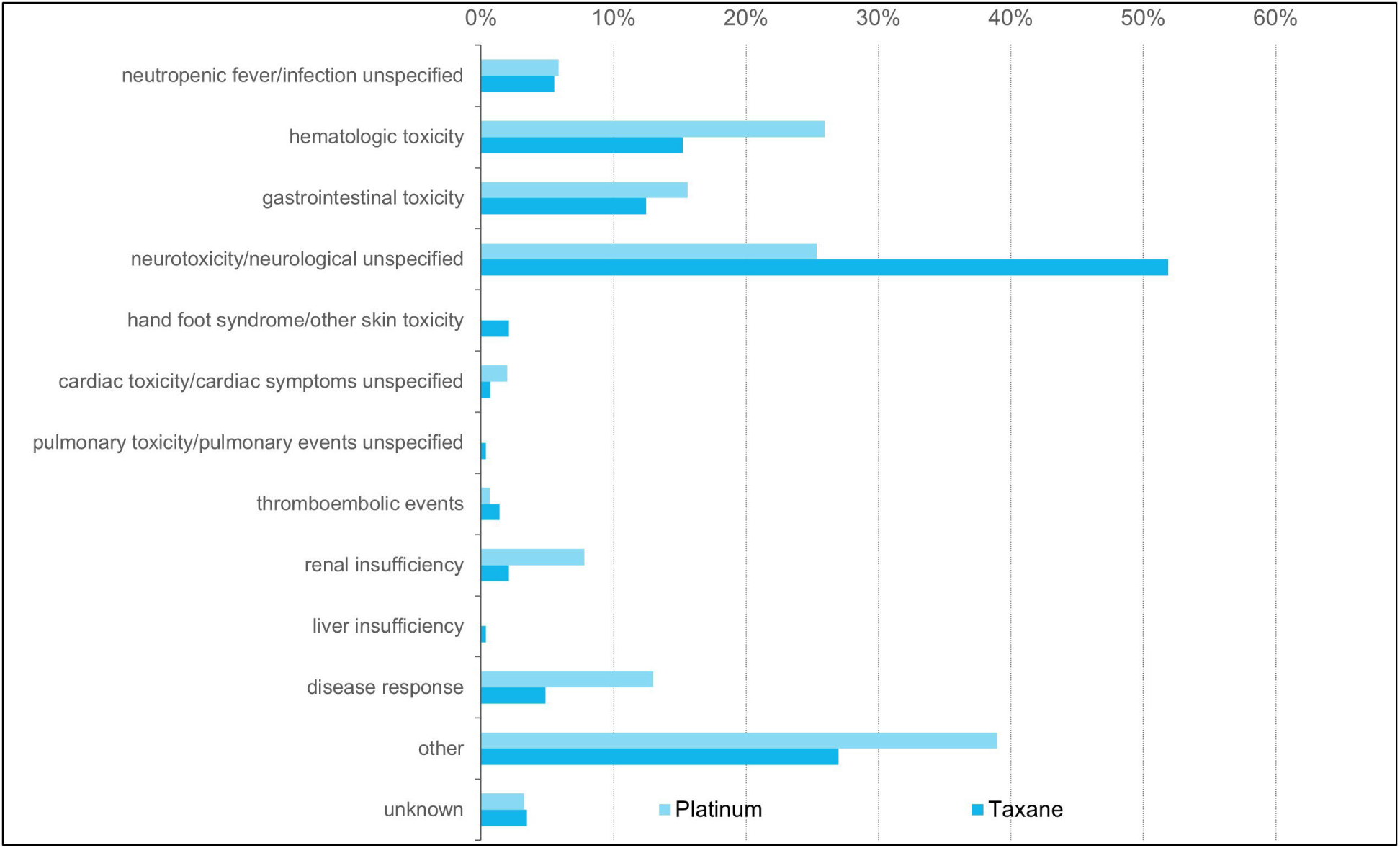
Reasons for reduction in the number of chemotherapy cycles in the entire cohort.

**Supplementary Figure 7.**
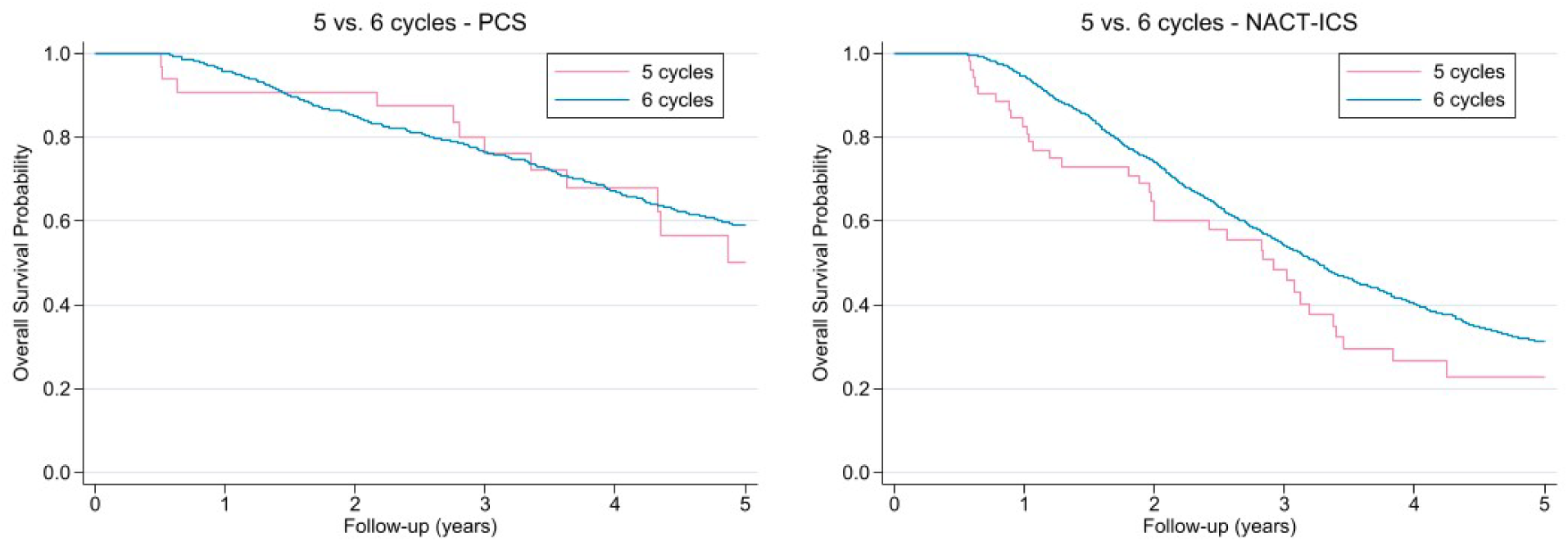
Kaplan-Meier survival curves for the patients who underwent 5 vs. 6 chemotherapy cycles in the primary cytoreductive surgery and interval cytoreductive surgery setting. *Hazard ratios (HR) from multivariable Cox proportional hazards models: primary cytoreductive surgery setting (HR 1.53, 95%–CI: 0.84–2.76.19); interval cytoreductive surgery setting (HR 1.25, 95%–CI: 0.88–1.76).

## Notes

### Competing Interest Statement

The authors have declared no competing interest.

### Author Declarations

The scientific committee of the Pillar Oncology of the Dutch Society of Obstetrics and Gynaecology (NVOG) and the Privacy Review Board of Integraal Kankercentrum Nederland (IKNL) gave approval to this study (K23.306; 23-11-2023). The institutional review board waived the need for informed consent because of the study's retrospective design.

